# Longitudinal Variability of Lipoprotein(a) in Children with Type 1 Diabetes: Implications for Cardiovascular Risk Stratification

**DOI:** 10.64898/2026.03.28.26349624

**Authors:** Fanny Iafrate-Luterbacher, Cecilia Jimenez-Sanchez, Maria Loukia Anastasiadou, Julien Prados, Frida Renström, Michael Brändle, Stefan Bilz, Valerie M Schwitzgebel

## Abstract

**Context:** Lipoprotein(a) [Lp(a)] is a genetically determined and independent cardiovascular risk factor, traditionally considered stable across the lifespan, supporting a single lifetime measurement strategy. However, its longitudinal behavior during childhood and adolescence remains poorly characterized, particularly in individuals with type 1 diabetes who are at increased lifetime risk of cardiovascular disease.

**Objective:** We aimed to characterize intra- and inter-individual trajectories of Lp(a) in youth with type 1 diabetes and to assess the implications of variability for cardiovascular risk classification.

**Methods:** We conducted a retrospective single-center cohort study of children and adolescents with type 1 diabetes followed at Geneva University Hospitals between 2012 and 2023. Annual fasting Lp(a) concentrations were analyzed longitudinally. Variability was assessed in participants with more than two measurements. Clinically relevant thresholds were used to evaluate risk reclassification. Statistical analyses included paired Wilcoxon tests, Pearson and Kendall correlations, and Holm-adjusted p-values.

**Results:** A total of 287 participants contributed 1,408 Lp(a) measurements over a median follow-up of 6.2 years (IQR 2.9–9.6). At baseline, 26% had elevated Lp(a) (above or equal 300 mg/L). Among participants with serial measurements, 32% exhibited intraindividual fluctuations exceeding 50% of their maximum value. Reclassification across the 300 mg/L threshold occurred in 11.9% of participants. Lp(a) concentrations peaked between ages 10 and 13 years and declined thereafter. Modest seasonal variation was observed, with higher levels in autumn and winter (P < 0.05).

**Conclusions:** In youth with type 1 diabetes, Lp(a) demonstrates clinically relevant intraindividual variability over time. These findings suggest that reliance on a single lifetime measurement may lead to misclassification of cardiovascular risk and support repeated assessment, particularly during adolescence, to improve risk stratification.

## Introduction

Lipoprotein(a) [Lp(a)] is a genetically determined and independent risk factor for cardiovascular disease (CVD). Its concentration is largely governed by variants in the *LPA* gene and has traditionally been considered stable across the lifespan, supporting the recommendation of a single lifetime measurement for cardiovascular risk assessment (1,2). However, this assumption is based primarily on adult data, and longitudinal evidence in pediatric populations remains limited.

Children with early-onset type 1 diabetes are at increased lifetime risk of CVD, with coronary artery disease developing earlier than in the general population (3). Subclinical vascular abnormalities, including dyslipidemia, inflammation, and endothelial dysfunction, often emerge during childhood and adolescence, contributing to long-term cardiovascular risk (3). In this context, accurate identification of lipid-related risk factors early in life is essential.

Lp(a) has attracted increasing attention as a causal and independent contributor to atherogenesis, also in youth (4,5). Structurally analogous to low-density lipoprotein, it consists of an apolipoprotein B-100 particle covalently linked to apolipoprotein(a), which carries proatherogenic oxidized phospholipids (6). Elevated Lp(a) promotes vascular disease through pro-inflammatory, pro-thrombotic, and antifibrinolytic mechanisms (7), and genetic studies have identified *LPA* variants as major determinants of coronary artery disease risk (8). Clinically, concentrations above 300 mg/L are associated with increased cardiovascular risk (1).

Emerging therapies targeting Lp(a), including antisense oligonucleotides and small interfering RNAs, have demonstrated substantial and sustained reductions in circulating levels in adults (9), underscoring the growing importance of identifying at-risk individuals early in life.

Despite its strong genetic determination, it remains unclear whether Lp(a) behaves as a stable biomarker during childhood and adolescence, particularly in high-risk populations such as individuals with type 1 diabetes. Clarifying its longitudinal behavior is essential to determine whether a single measurement is sufficient for risk stratification.

We therefore aimed to characterize intra- and inter-individual trajectories of Lp(a) in a longitudinal cohort of children and adolescents with type 1 diabetes and to evaluate the implications of its variability for cardiovascular risk classification.

## Methods

We conducted a retrospective, single-center study at Geneva University Hospitals analyzing fasting Lp(a) annually from 2012 to 2023 (n = 1,408 measurements). To be eligible, patients had to meet the following criteria: diagnosis of type 1 diabetes, at least one Lp(a) measurement, a minimum of one year of follow-up by the Geneva pediatric diabetes unit, and age under 25 years. Yearly blood analyses were performed in the morning after an overnight fast. Annual blood draws were performed only when participants were clinically well and postponed in case of acute illness (e.g., fever or viral infection). Data on demographic characteristics (age, sex, BMI), insulin dose, and laboratory measurements including Lp(a), hemoglobin A1c, triglycerides, total cholesterol, HDL cholesterol, and LDL cholesterol were collected at diagnosis and after each annual clinical assessment. The REDCap platform was used for data collection (10). Lp(a) measurements were performed using an immunoturbidimetric method with a coefficient of variation below 8.3% (Roche Diagnostics SA No art. 05852625 190, instrument: Roche Cobas 8000), which is isoform insensitive.

### Statistical Methods

We performed descriptive analyses for all study variables. For continuous variables, means, SDs, and medians were calculated; frequency counts and percentages were calculated for categorical variables. To assess the association between Lp(a) levels and predefined factors (BMI, ethnicity, glycated hemoglobin, daily insulin dose, sex, age), we used two-sample Wilcoxon tests for categorical variables and Kendall or Pearson correlation tests for continuous variables, depending on data distribution. When appropriate, p-values were adjusted for multiple testing using Holm’s method. Statistical significance was set at P < 0.05 for all analyses. To analyze intraindividual variability in Lp(a) concentrations over time, we restricted the analysis to 236 patients with at least two annual assessments. Changes in Lp(a) levels between consecutive visits were quantified. For the seasonality analysis, changes in Lp(a) were calculated relative to each patient’s median Lp(a) level across visits. All preprocessing, analysis, and visualization were performed using the R programming language [R: A Language and Environment for Statistical Computing; R Core Team; https://www.R-project.org/, 2025].

### Risk curves

To estimate the risk for a patient to reach a pathological Lp(a) levels of 300mg/L (resp. 500mg/L and 1000mg/L) given an observed Lp(a) level x, we computed from our data the ratio M(x)/N(x), where N(x) is the number of patient whose range of variation of Lp(a) include the value x, while M(x) is the number of patient whose range of variation of Lp(a) include both x and 300 (resp. 500 and 1000).

## Results

Of 435 eligible patients, 287 children and adolescents with type 1 diabetes were included (Fig. 1, 2A). The median follow-up was 6.2 years (IQR 2.9–9.6). Baseline characteristics are provided in Supplementary Table 1. Lp(a) concentrations were stratified into six categories based on established coronary artery disease risk thresholds (Fig. 2B), with ≥300 mg/L indicating elevated cardiovascular risk (2).

**Fig. 1.**
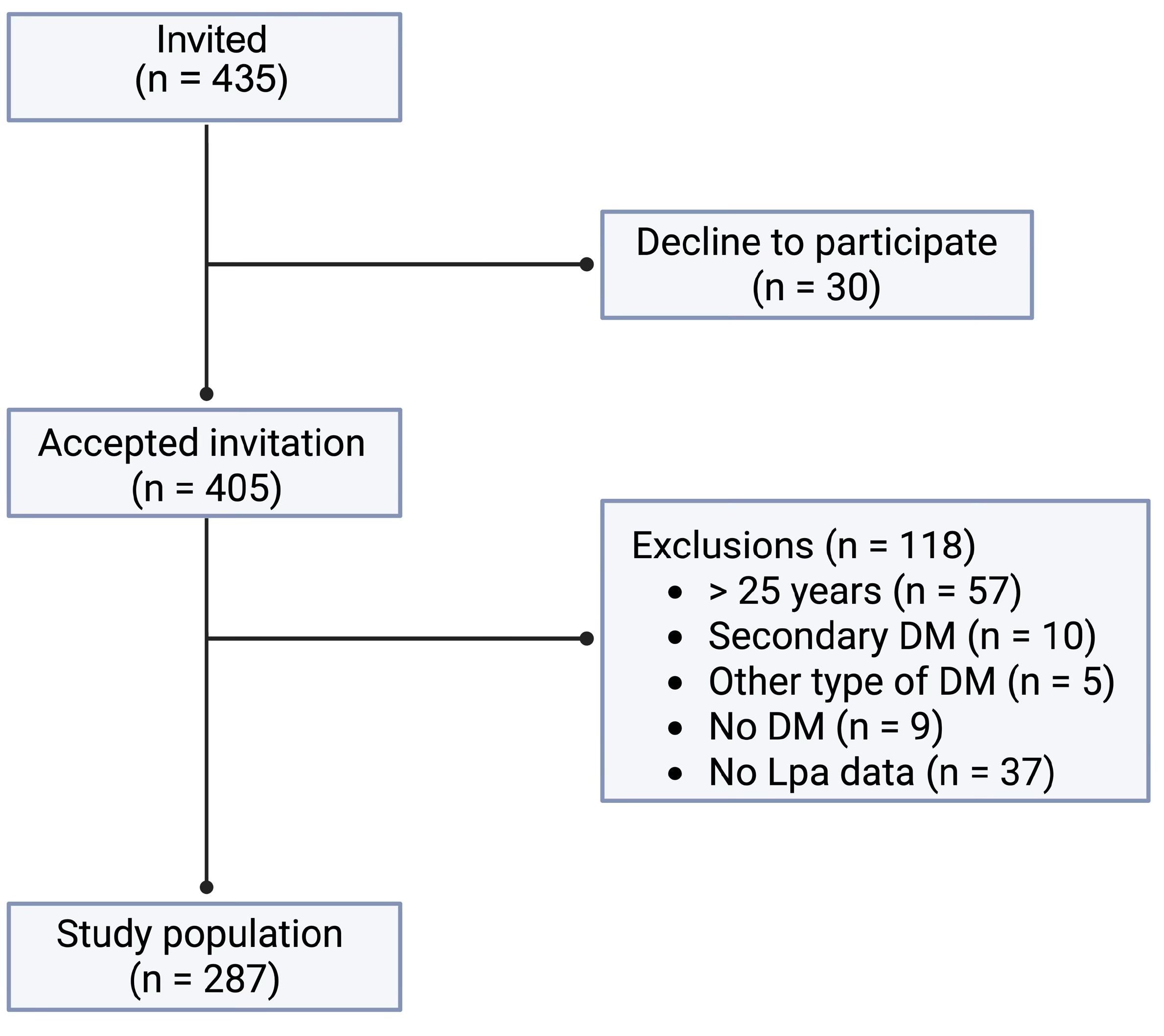
Flowchart of recruitment of participants. DM: Diabetes mellitus

**Fig. 2.**
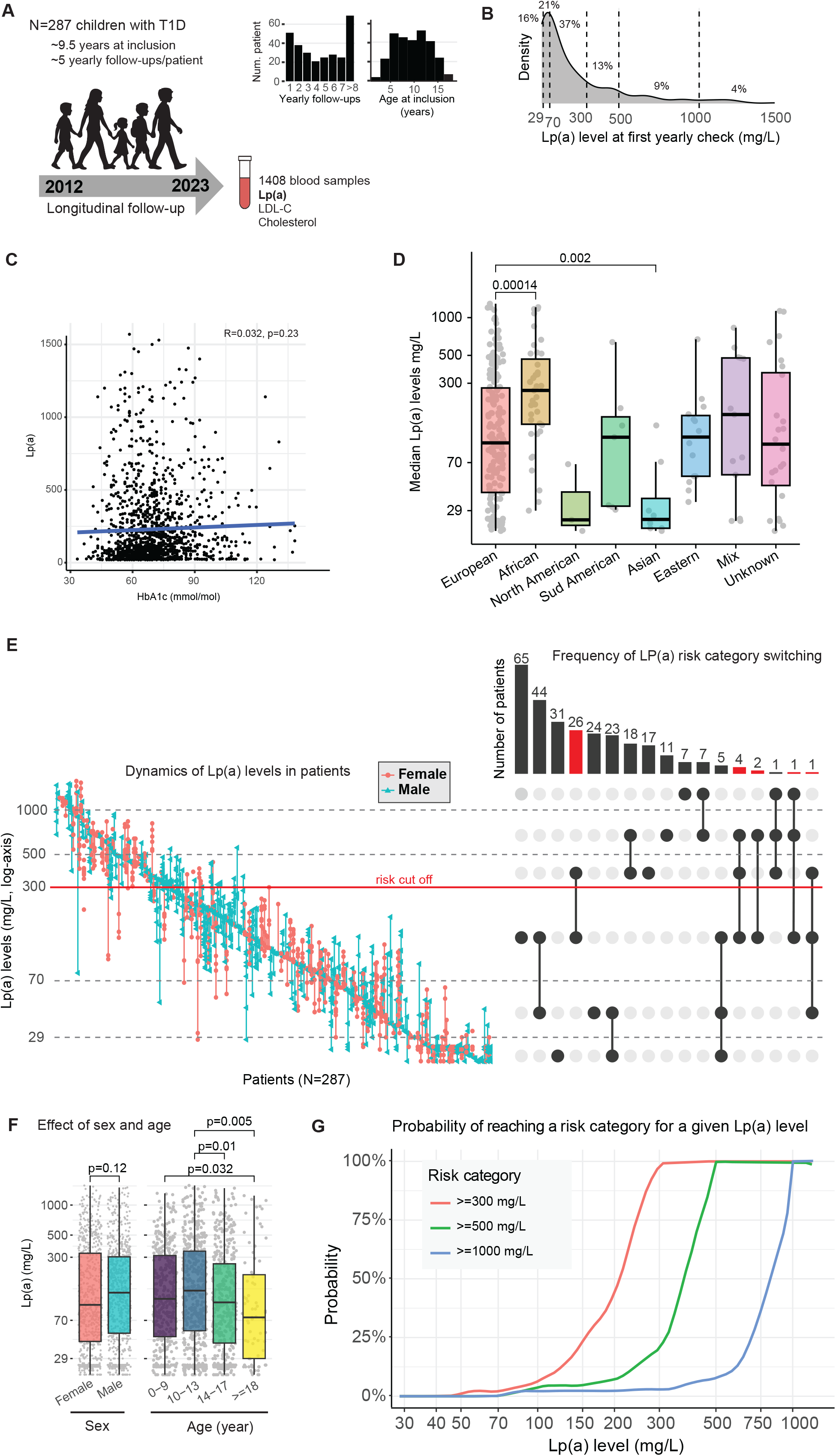
Longitudinal variability and risk stratification of lipoprotein(a) in youth with type 1 diabetes. **A)** Overview of cohort follow-up and sampling. Left: A total of 287 children with type 1 diabetes were followed longitudinally for up to 11 years, yielding 1408 fasting blood samples. Middle: Median age at inclusion was 9.5 years: Right: Patients underwent a mean of 5 annual follow-up visits. **B)** Density distribution of Lp(a) levels at first yearly check, with dashed lines indicating the five standard diagnostic thresholds for Lp(a) and labels showing the percentage of patients within each category. **C)** Correlation between Lp(a) and HbA1c levels (participant median values). **D)** Median Lp(a) levels according to patient ancestry. The ancestry composition of the cohort was as follows: 179 (62.4%) European, 39 (13.6%) African, 14 (4.9%) Middle Eastern, 8 (2.8%) East Asian, 7 (2.4%) South American, 3 (1.0%) North American, 13 (4.5%) mixed ancestry, and 24 (8.4%) unknown ancestry. Pairwise comparisons were performed using Wilcoxon tests with adjustment for multiple testing; only adjusted P-values ≤ 0.05 are shown. **E)** Longitudinal variability of Lp(a) and risk category transitions. Left: Individual patient trajectories of Lp(a) levels across all visits, sorted relatively to each patient’s median Lp(a) value: females (red points) and males (green triangles). Right: UpSet plot summarizing the number of patients transitioning between Lp(a) risk categories over time (in red). **F)** Age- and sex-stratified Lp(a) levels. Boxplots of all 1,408 Lp(a) measurements stratified by patient sex and age group at the time of sampling. No significant difference was observed between females and males (P = 0.12). Repeated measurements from the same participant are included (mean ≈ 4.9 measurements per patient). Statistical comparisons between age groups were performed using Wilcoxon tests with correction for multiple testing; only adjusted P-values ≤ 0.05 are shown as significant. **G)** Modelled probabilities of crossing Lp(a) risk thresholds. Risk curves depicting the estimated probability that an individual patient’s Lp(a) measurement will exceed clinically relevant thresholds of 300 mg/L, 500 mg/L, or 1,000 mg/L, based on baseline Lp(a) values. This modelling provides a dynamic tool for cardiovascular risk assessment over time.

At baseline, 26% of participants had elevated Lp(a) (≥300 mg/L), including 4% with levels exceeding 1,000 mg/L. Thirty-seven percent had low (<70 mg/L) or undetectable (<29 mg/L) concentrations, while 37% were within the intermediate range (70–<300 mg/L) (Fig. 2B). Lp(a) concentrations were not associated with HbA1c (Fig. 2C), sex, age at diabetes onset, BMI, or insulin dose (Supplementary Table 1). However, levels differed by ancestry, with higher concentrations observed in participants of African descent and lower concentrations in those of East Asian ancestry compared with European participants (Fig. 2D).

Among 236 participants with at least two measurements, 32% exhibited intraindividual fluctuations exceeding 50% of their individual maximum value. Variability was both increasing and decreasing across consecutive visits, without a consistent directional trend. Reclassification across the 300 mg/L threshold occurred in 11.9% of participants (Fig. 2E). Specifically, 9% transitioned between normal and elevated risk, and 2.4% shifted between low and very high risk (>1,000 mg/L). Variability was observed in both sexes.

Lp(a) concentrations followed a distinct age-related trajectory, with a peak between 10 and 13 years and a subsequent decline during late adolescence (Fig. 2F). We further estimated the probability of exceeding clinically relevant thresholds (300, 500, and 1,000 mg/L) according to baseline Lp(a) levels, providing a framework for dynamic risk stratification (Fig. 2G).

At the first annual assessment, 78% of participants had no fasting dyslipidemia. Elevated LDL-C (>3.4 mmol/L) was observed in 10%, including 1.7% with levels >5 mmol/L (Fig. 3A). LDL-C variability was present in both sexes (Fig. 3B). Notably, 9% of participants had concomitant elevations in Lp(a) and LDL-C, a lipid profile associated with increased cardiovascular risk (11).

**Fig. 3.**
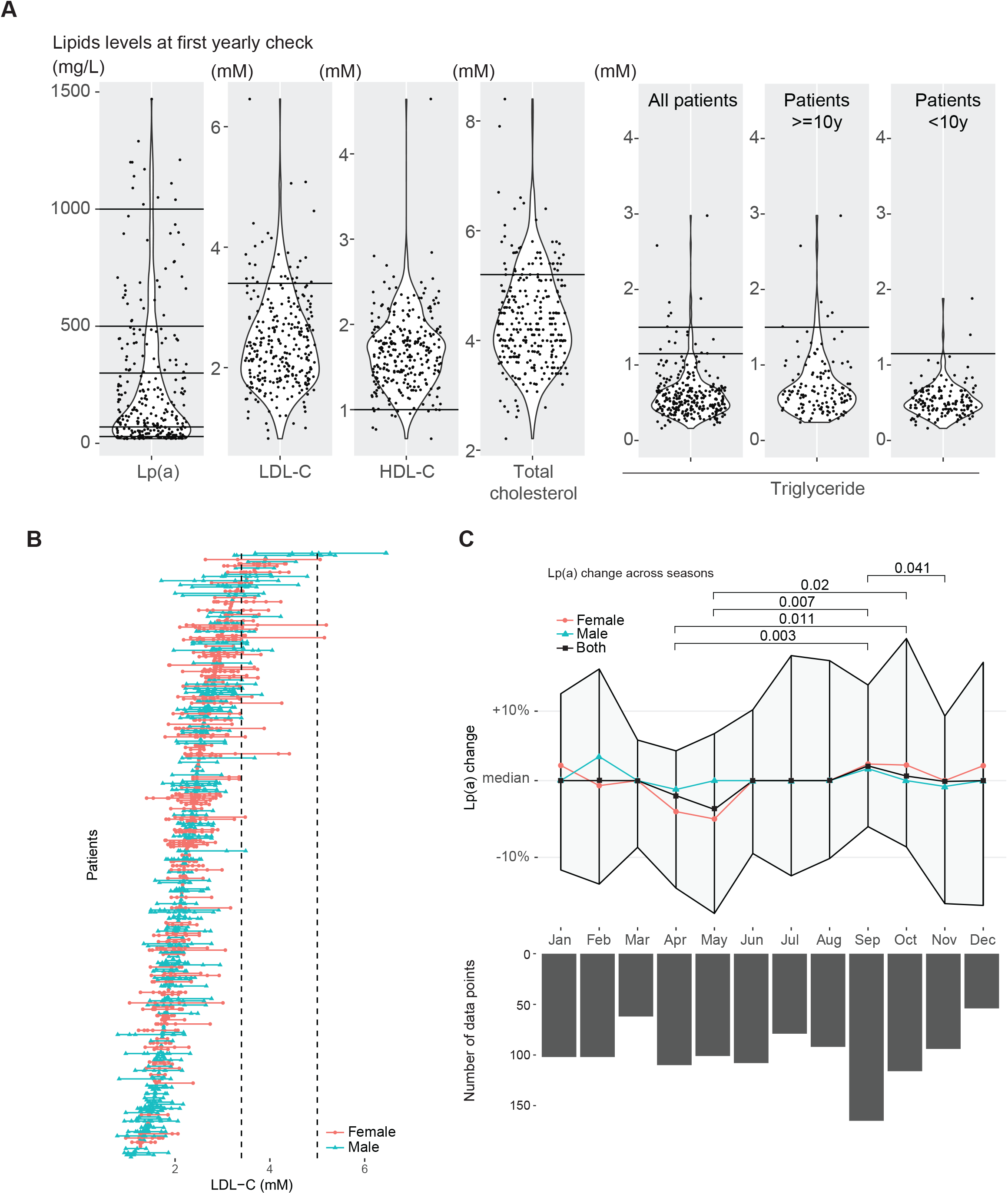
Lipid profiles and seasonal variation of lipoprotein(a) in youth with type 1 diabetes. **A)** Lipid profiles at first annual assessment. Violin plots of Lp(a) and lipid parameters measured at the first annual visit. Horizontal lines indicate clinically relevant thresholds: Lp(a): >1000 mg/L, >500–1000 mg/L, 300–500 mg/L, 70– <300 mg/L, 29–<70 mg/L, and <29 mg/L; LDL-C: 3.4 mmol/L; HDL-C: 1 mmol/L; total cholesterol: 5.2 mmol/L; triglycerides: 1.12 mmol/L and 1.46 mmol/L. Triglycerides are stratified by age due to age-specific reference ranges. **B)** Distribution of LDL-C levels across the cohort. LDL-C levels across all participants, ordered according to each patient’s median LDL-C value over time. **C)** Seasonal variation in Lp(a) levels. Seasonal variation of Lp(a) levels based on 1,185 measurements from 168 patients with at least four Lp(a) assessments. The y-axis represents the percent change in Lp(a) relative to each patient’s median value across all visits. Statistical significance between months was assessed using Wilcoxon tests with adjustment for multiple testing. The black line represents the median change across all patients, while red and blue lines indicate sex-specific trends. Shaded areas represent interquartile ranges. The number of observations per month is indicated.

In 268 participants with at least four Lp(a) measurements (n = 1,185), seasonal variation was observed. Lp(a) concentrations were modestly but significantly higher during autumn and winter compared with spring and summer, with the largest differences between April and September (Fig. 3C). Changes were calculated relative to each individual’s median Lp(a) across visits.

## Discussion

In this longitudinal cohort of children and adolescents with type 1 diabetes, we show that Lp(a), traditionally considered a genetically determined and stable biomarker, exhibits substantial intraindividual variability across childhood and adolescence. Notably, more than 11% of participants crossed clinically relevant cardiovascular risk thresholds over time, indicating that reliance on a single measurement may lead to risk misclassification.

Beyond its descriptive relevance, this variability may have biological and clinical implications. Analogous to glycemic or lipid variability, fluctuations in Lp(a) could contribute to vascular injury through intermittent exposure to proatherogenic lipoprotein profiles, potentially promoting oxidative stress, endothelial dysfunction, and low-grade inflammation key processes in early atherogenesis (12,13).

We identified a distinct age-related trajectory, with Lp(a) levels peaking between 10 and 13 years and declining thereafter. This pattern suggests a potential influence of pubertal hormonal changes, including sex steroids and growth factor signaling, on hepatic Lp(a) production (14). Adolescence may therefore represent a clinically relevant window during which Lp(a)-based cardiovascular risk assessment is particularly informative.

Seasonal variation further supports the concept that Lp(a) is not entirely biologically invariant. Higher concentrations during autumn and winter may reflect inflammatory modulation, potentially mediated by cytokines such as interleukin-6, as well as environmental influences including vitamin D status and dietary factors (14,15). Although modest in magnitude, these differences suggest that both developmental and environmental factors may influence Lp(a) concentrations.

A subset of participants exhibited concomitant elevations in Lp(a) and LDL-C, a lipid profile associated with increased cardiovascular risk (11). Because Lp(a)-associated cholesterol contributes to measured LDL-C levels, this overlap may complicate risk assessment and potentially obscure conditions such as familial hypercholesterolemia (16,17). These findings support the inclusion of Lp(a) in lipid evaluation in pediatric diabetes populations.

The ancestry-related differences observed in our cohort are consistent with previous reports and underscore the importance of considering population-specific distributions in cardiovascular risk assessment (18,19).

Taken together, our findings indicate that Lp(a) may not behave as a strictly static biomarker during childhood and adolescence but instead demonstrates clinically relevant variability. These observations support the potential value of longitudinal assessment, particularly during key developmental stages.

### Strengths and limitations

This study has several strengths. Its longitudinal design, with repeated measurements over a median follow-up exceeding six years, enabled robust assessment of intraindividual variability across key developmental stages. The use of an isoform-insensitive assay enhances the reliability of Lp(a) quantification, and the detailed clinical characterization allowed evaluation of relevant metabolic and demographic factors.

Limitations should also be considered. The single-centre, retrospective design may limit generalisability to other populations. The absence of genetic data on *LPA* variants and inflammatory markers precludes mechanistic insights into the determinants of Lp(a) variability. In addition, the study was not designed to assess clinical cardiovascular outcomes.

## Conclusions

Lp(a) demonstrates clinically meaningful intraindividual variability in children and adolescents with type 1 diabetes, resulting in reclassification of cardiovascular risk in a significant subset of patients. These findings suggest that a single lifetime measurement may be insufficient and support repeated assessment, particularly during adolescence, to improve risk stratification. A longitudinal approach to Lp(a) monitoring may facilitate earlier identification of high-risk individuals and inform preventive strategies in this population.

## Data Availability

Deidentified data are available from the corresponding author upon reasonable request and with appropriate justification, in accordance with institutional and ethical regulations.

## Abbreviations

CVD: Cardiovascular disease
HDL-C: High-density lipoprotein cholesterol
LDL-C: Low-density lipoprotein cholesterol
Lp(a): Lipoprotein (a)

## Declarations

### Ethics approval and consent to participate

This study was approved by the Geneva Ethics Committee (CCER 2022-02251) and conducted in accordance with the principles of the Declaration of Helsinki. All patients included in the study were informed about the use of their data and were given the opportunity to decline participation. Informed consent for the use of clinical data for research purposes, as well as for publication and future research, was obtained in accordance with institutional and national regulations.

### Consent for publication

Not applicable.

### Authors’ contributions

F.I.L. contributed to the conception and design of the study, data collection, database creation, and co-wrote the manuscript. C.J.S. and M.A. contributed to data collection, database creation, and critical revision of the manuscript. J.P. performed the statistical analyses and critically reviewed the manuscript. F.R., M.B., and S.B. contributed to manuscript editing and critical revision. V.M.S. contributed to the conception and design of the study, participated in data interpretation and discussion, and co-wrote, edited, and critically revised the manuscript. V.M.S. is the guarantor of this work and, as such, had full access to all the data in the study and takes responsibility for the integrity of the data and the accuracy of the data analysis. All authors read and approved the final manuscript.

## Acknowledgements

We thank the Clinical Pediatric Research Platform of the Children’s University Hospital of Geneva for their assistance, as well as all the families and children who participated in this project.

## Additional file

**Supplementary Table 1.**
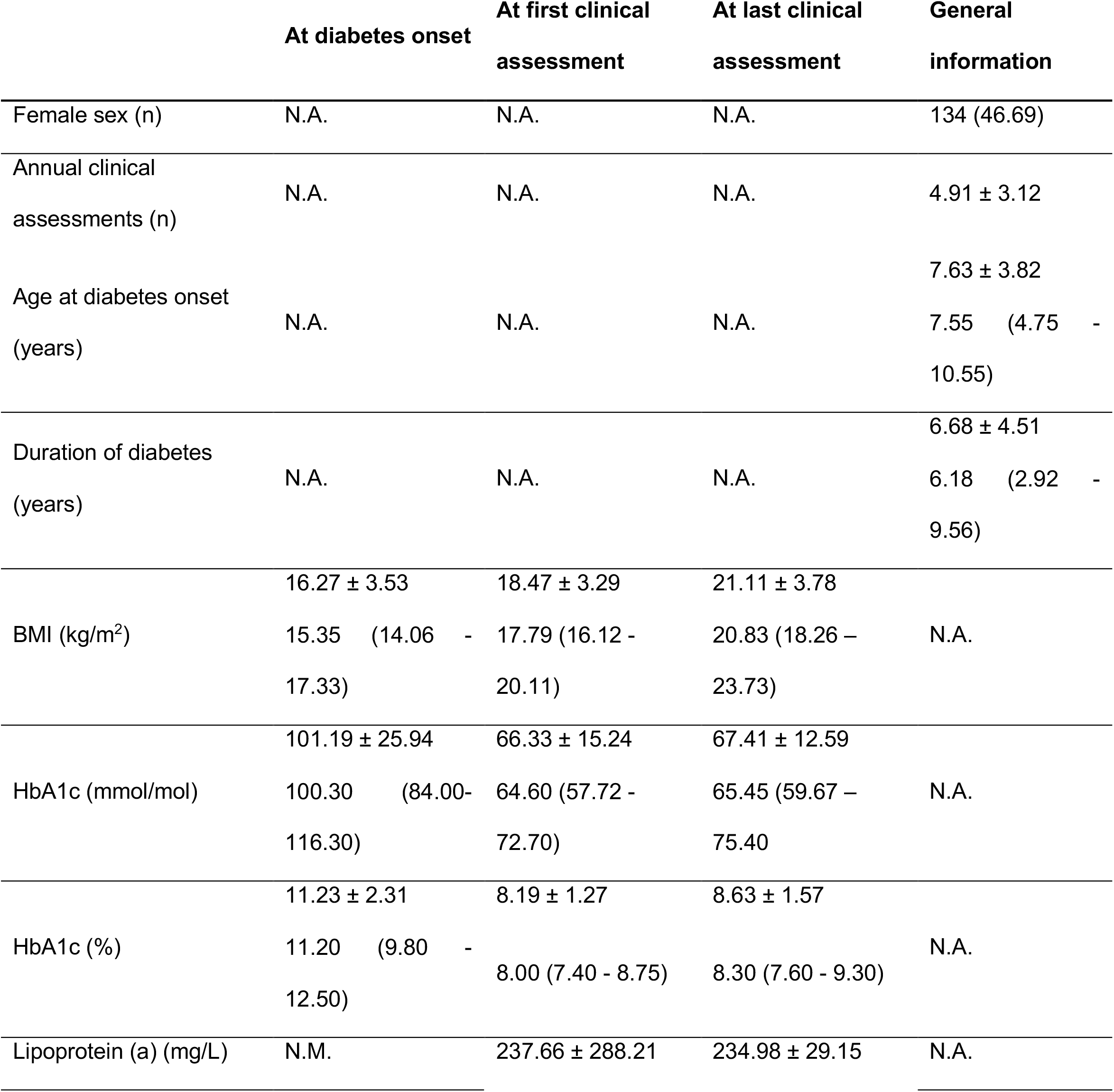
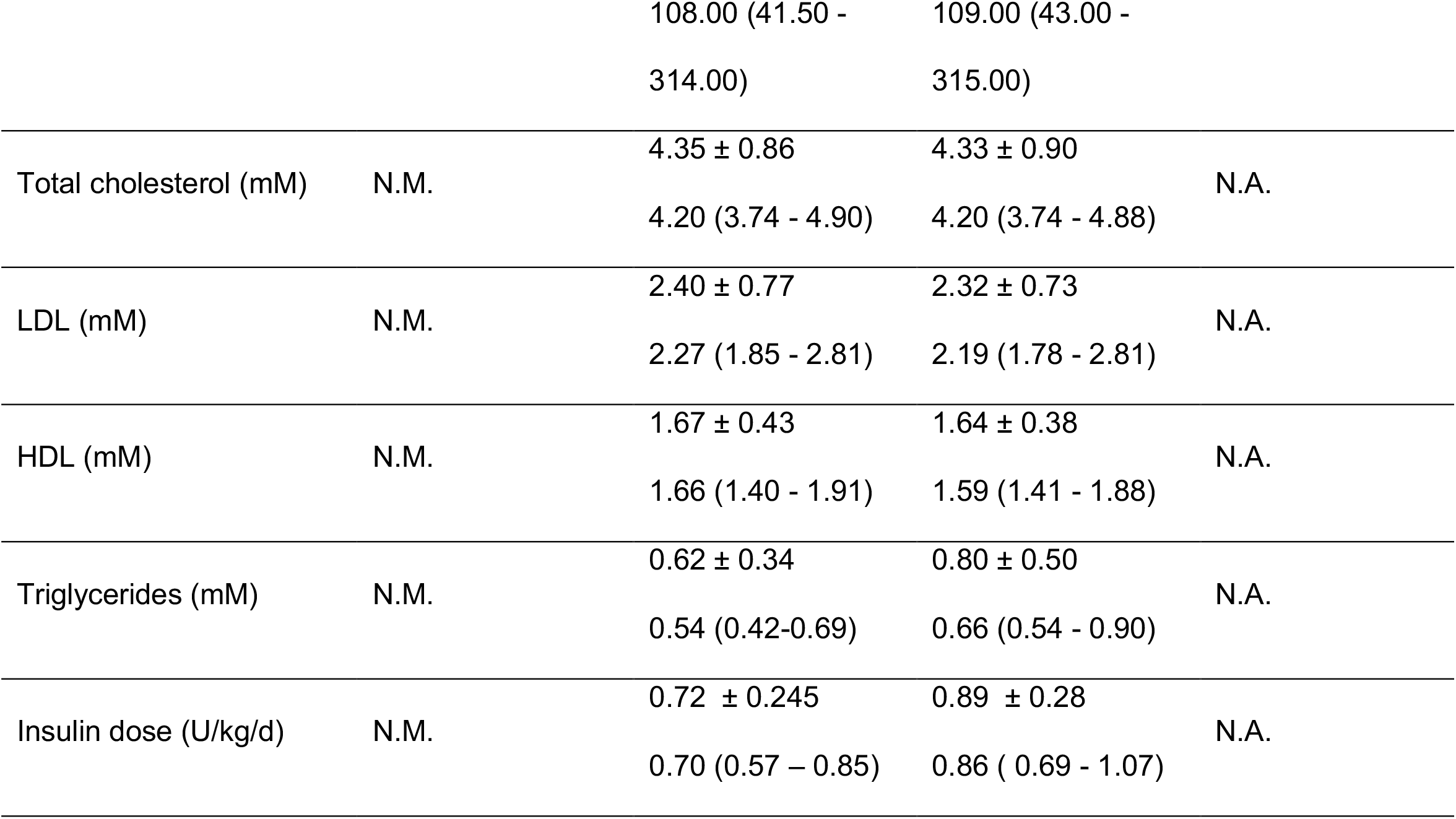
Characteristics of study participants. Case subjects n=287. Results are expressed in mean ± SD and/or median and IQR (25^th –^ 75^th^), except for sex, which is expressed as n (%). Duration of diabetes was assessed at the last clinical assessment. N.M. = not measured, N.A. = not applicable.

|  | At diabetes onset | At first clinical assessment | At last clinical assessment | General information |
| --- | --- | --- | --- | --- |
| Female sex (n) | N.A. | N.A. | N.A. | 134 (46.69) |
| Annual clinical assessments (n) | N.A. | N.A. | N.A. | 4.91 ± 3.12 |
| Age at diabetes onset (years) | N.A. | N.A. | N.A. | 7.63 ± 3.82<br>7.55 (4.75 - 10.55) |
| Duration of diabetes (years) | N.A. | N.A. | N.A. | 6.68 ± 4.51<br>6.18 (2.92 - 9.56) |
| BMI (kg/m<sup>2</sup>) | 16.27 ± 3.53<br>15.35 (14.06 - 17.33) | 18.47 ± 3.29<br>17.79 (16.12 - 20.11) | 21.11 ± 3.78<br>20.83 (18.26 – 23.73) | N.A. |
| HbA1c (mmol/mol) | 101.19 ± 25.94<br>100.30 (84.00- 116.30) | 66.33 ± 15.24<br>64.60 (57.72 - 72.70) | 67.41 ± 12.59<br>65.45 (59.67 – 75.40) | N.A. |
| HbA1c (%) | 11.23 ± 2.31<br>11.20 (9.80 - 12.50) | 8.19 ± 1.27<br>8.00 (7.40 - 8.75) | 8.63 ± 1.57<br>8.30 (7.60 - 9.30) | N.A. |
| Lipoprotein (a) (mg/L) | N.M. | 237.66 ± 288.21 | 234.98 ± 29.15 | N.A. |
|  |  | 108.00 (41.50 -<br>314.00) | 109.00 (43.00 -<br>315.00) |  |
| Total cholesterol (mM) | N.M. | 4.35 ± 0.86<br>4.20 (3.74 - 4.90) | 4.33 ± 0.90<br>4.20 (3.74 - 4.88) | N.A. |
| LDL (mM) | N.M. | 2.40 ± 0.77<br>2.27 (1.85 - 2.81) | 2.32 ± 0.73<br>2.19 (1.78 - 2.81) | N.A. |
| HDL (mM) | N.M. | 1.67 ± 0.43<br>1.66 (1.40 - 1.91) | 1.64 ± 0.38<br>1.59 (1.41 - 1.88) | N.A. |
| Triglycerides (mM) | N.M. | 0.62 ± 0.34<br>0.54 (0.42-0.69) | 0.80 ± 0.50<br>0.66 (0.54 - 0.90) | N.A. |
| Insulin dose (U/kg/d) | N.M. | 0.72 ± 0.245<br>0.70 (0.57 – 0.85) | 0.89 ± 0.28<br>0.86 ( 0.69 - 1.07) | N.A. |

